# An exploratory study of urinary proteome in trigeminal neuralgia

**DOI:** 10.1101/2024.12.02.24318147

**Authors:** Lilong Wei, Haitong Wang, Yun Zhou, Jianqiang Wu, Yuliang Zhan, Yongtong Cao, Youhe Gao

## Abstract

Trigeminal neuralgia (TN) is a neurological disorder characterized by severe pain, with a complex pathogenesis that seriously affects the quality of life of patients. Urinary proteomics has shown great potential in disease research. The aim of this study is to explore the diagnostic and therapeutic value of urine proteomics in TN. The urine of 10 cases of TN and 11 healthy individuals were studied using liquid chromatography-mass spectrometry (LC-MS) proteomics technology. Group analysis and one to many individual analysis strategies were used in quantitative analysis. A total of 2620 proteins were identified in group analysis, of which 1865 were quantifiable proteins. The majority of specimens in each group could be distinguished by unsupervised clustering analysis of urine proteome. The biological processes of immune response regulation signal pathway, immune response, natural killer cell activation, fatty acid transport, and natural killer cell inhibition signal pathway, as well as the KEGG pathway of antigen processing and presentation and natural killer cell-mediated cytotoxicity, were significantly enriched in the comparison between the two populations. An individual analysis revealed that there were more significantly different proteins among individuals, with 15 proteins identified in at least 9 different patients. The urinary proteome provided molecular characteristics of urinary proteins for patients with TN, describing the changes that occur in the patient’s body. The strong enrichment of its immune response might be related to the onset of the disease, providing new directions for immune related treatment of the disease; the significant differential proteins could become potential disease markers for disease diagnosis and treatment evaluation.

## 1 Introduction

Trigeminal neuralgia (TN) is a type of neurological pain characterized by severe facial pain, usually unilateral in nature. The pain is typically paroxysmal, stabbing, or electric shock-like, lasting from a few seconds to a few minutes, and occurs at a relatively high frequency^1^. TN often affects the condition of basic human psychological, physiological and social needs and activities, such as touching the face, speaking, eating and drinking^2^. The symptoms of TN are obvious, however, there is currently no gold standard or specific method or biomarker to record it^3^. The development of proteomics technology tends to comprehensively discover disease markers^4,5^, urine, as the most non-invasive source of disease markers, is widely used in clinical research^6,7^,at present, there is no search for the use of urinary proteomics technology to study related diseases. This study is the first attempt to use urinary proteomics to study TN, aiming to explore the characteristics of urinary proteomics in patients with TN and to find urine nerve injury related markers.

## 2 Materials and methods

### 2.1 Experimental design and urine sample collection

This study was approved by the Ethics Committee of China-Japan Friendship Hospital (Approval No. 2023-ky-126), and participants were from patients and healthy physical examinees of China-Japan Friendship Hospital. Inclusion criteria for participants: The diagnosis of TN was clear; Exclusion criteria: Tumor diseases were excluded; kidney diseases were excluded; Healthy people were excluded from nerve-related diseases, tumor diseases, kidney diseases. Urine proteomics technology was used to analyze healthy people and patients with TN (Fig. 1, Experimental route). The participant samples included 11 urine samples from healthy individuals (N) and 10 urine samples from patients with TN. All samples were collected after clinical testing and stored in a -80°C refrigerator.

**Figure 1.**
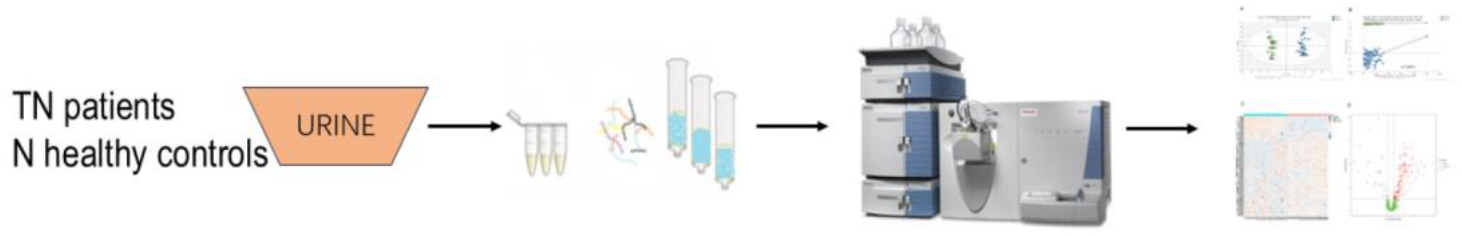
Research flow chart of quantitative urinary proteomics study in TN and N

### 2.2 Processing of urine samples

Extraction and quantification of urinary proteins: The collected urine samples were centrifuged at 12000×g for 30 min at 4°C, and the supernatant was transferred to a 50 mL centrifugal tube. Dithiothreitol solution (DTT, Sigma) was then added to a final concentration of 20 mM, and the mixture was shaken and incubated in a water bath at 37°C for 1 h before being cooled to room temperature. Iodoacetamide (IAA, Sigma) was added to a final concentration of 50 mM, and the mixture was shaken and reacted in the dark at room temperature for 40 min. Six times the volume of pre-cooled absolute ethanol was added, and the mixture was homogeneously mixed and precipitated at -20°C for 24 h. On the second day, the mixture was centrifuged at 4°C, 12000×g for 30 min, and the supernatant was discarded. The protein precipitate was resuspended in lysis buffer (containing 8 mol/L urea, 2 mol/L thiourea, 25 mmol/L dithiothreitol, 50 mmol/L Tris). After centrifugation at 12000×g for 30 min at 4°C, the supernatant was placed in a new EP tube. The protein concentration was measured by the Bradford method. Urine protein digestion: 100 μg urine protein sample was added to the filter membrane of 10 kDa ultrafiltration tube (Pall, Port Washington, NY, USA) and placed in an EP tube, and 25 mmol/L NH_4_HCO_3_ solution was added to make the total volume 200 μL. Then the membrane washing operation was carried out: ➀200 μL UA solution (8 mol/L urea, 0.1 mol/L Tris-HCl, pH 8.5) was added and centrifuged and washed twice at 14000×g 5 min 18°C; ➁ Loading: the sample was added and centrifuged at 14000×g 40 min 18°C; ➂ 200 μL UA solution was added and centrifuged at 14000×g for 40 min at 18°C, repeated 2 times; ➃ 25 mmol/L NH_4_HCO_3_ solution was added and centrifuged at 14000×g 40 min 18°C for 3-4 times; ➄ Trypsin (Trypsin Gold, Promega, Fitchburg, WI, USA) was added at a ratio of 1:50 for digestion, and the water bath was kept at 37°C for 12-16 h. The next day, the peptide segment was collected by centrifugation at 13000×g 30 min 4°C, and passed through the HLB column (Waters, Milford, MA, USA) for desalting. Then the sample were dried using a vacuum dryer and stored at -80°C.

### 2.3 LC-MS/MS tandem mass spectrometry analysis

The digested samples were dissolved in 0.1% formic acid and quantified using the BCA kit. The peptide concentration was diluted to 0.5 μg/μL. Four μL of each sample was taken to prepare the mixed polypeptide sample, and the separation was performed using a high pH reversed phase peptide separation kit (Thermo Fisher Scientific) according to the instructions. Ten fractions were collected by centrifugation, and after drying using a vacuum dryer, they were resuspended in 0.1% formic acid. The iRT reagent (Biognosys, Switzerland) was added at a sample: iRT volume ratio of 10:1 to calibrate the retention time of the extracted peptide peaks. For analysis, 1 μg of peptides from each sample was taken, and mass spectrometry analysis and data acquisition were performed using an EASY-nLC1200 chromatography system (Thermo Fisher Scientific, USA) and an Orbitrap Fusion Lumos Tribrid mass spectrometer (Thermo Fisher Scientific, USA). In order to generate the spectral library, the separated 10 Fractions were analyzed by mass spectrometry in Data Dependent Acquisition (DDA) mode. The mass spectrometry data were collected in high sensitivity mode. A complete mass spectrometry scan was obtained in the range of 350-1500m/z with a resolution setting of 60,000. Individual samples were analyzed using the Data Independent Acquisition (DIA) mode. DIA acquisition was performed using a DIA method with 36 windows. After every 8 samples, a single DIA analysis of pooled peptides was performed as quality control.

### 2.4 Database searching and Label-free DIA quantification

The raw data (RAW files) acquired from LC-MS/MS were imported into Proteome Discoverer (version 2.1, Thermo Scientific) and searched against the SwissProt database (taxonomy: Homo; containing 20346 sequences). The iRT sequences were added to the database for comparison. The search results were then imported into Spectronaut Pulsar (Biognosys AG, Switzerland) for processing and analysis. The abundance of peptides was calculated by summing the peak areas of their respective fragment ions in MS_2_. The protein intensity was calculated by summing the abundances of their respective peptides.

### 2.5 Data analysis

Each sample was performed in triplicate, and the average value was used for statistical analysis. The identified proteins were compared, and differentially expressed proteins were screened. The loose screening criteria for differentially expressed proteins were: fold change (FC) ≥ 1.5 or ≤ 0.67, and P value < 0.05 by two-tailed unpaired t-test analysis. The strict screening criteria for differentially expressed proteins were: FC ≥ 2 or ≤ 0.5, and P value < 0.01 by two-tailed unpaired t-test analysis. Functional enrichment analysis of the screened differentially expressed proteins was performed using the Wukong platform (https://www.omicsolution.org/wkomic/main/), Uniprot website (https://www.uniprot.org/) and DAVID database (https://david.ncifcrf.gov/). The reported literature was retrieved from the Pubmed database (https://pubmed.ncbi.nlm.nih.gov) for functional analysis of differentially expressed proteins.

## 3 Experimental results

### 3.1 Urine proteome mass spectrometry identification and analysis

A total of 2620 protein-specific peptides were identified in 11 urine samples of healthy people (N) and 10 patients with trigeminal neuralgia (TN) by LC-MS/MS proteomics, with protein level FDR<1%. 1865 proteins (missing values less than 50% in quality control samples) can be used for quantitative analysis. The correlation of quality control samples is shown in Figure 1. PCA analysis and OPLS-DA analysis of the two groups of samples were shown in Figure 2.

**Figure 2.**
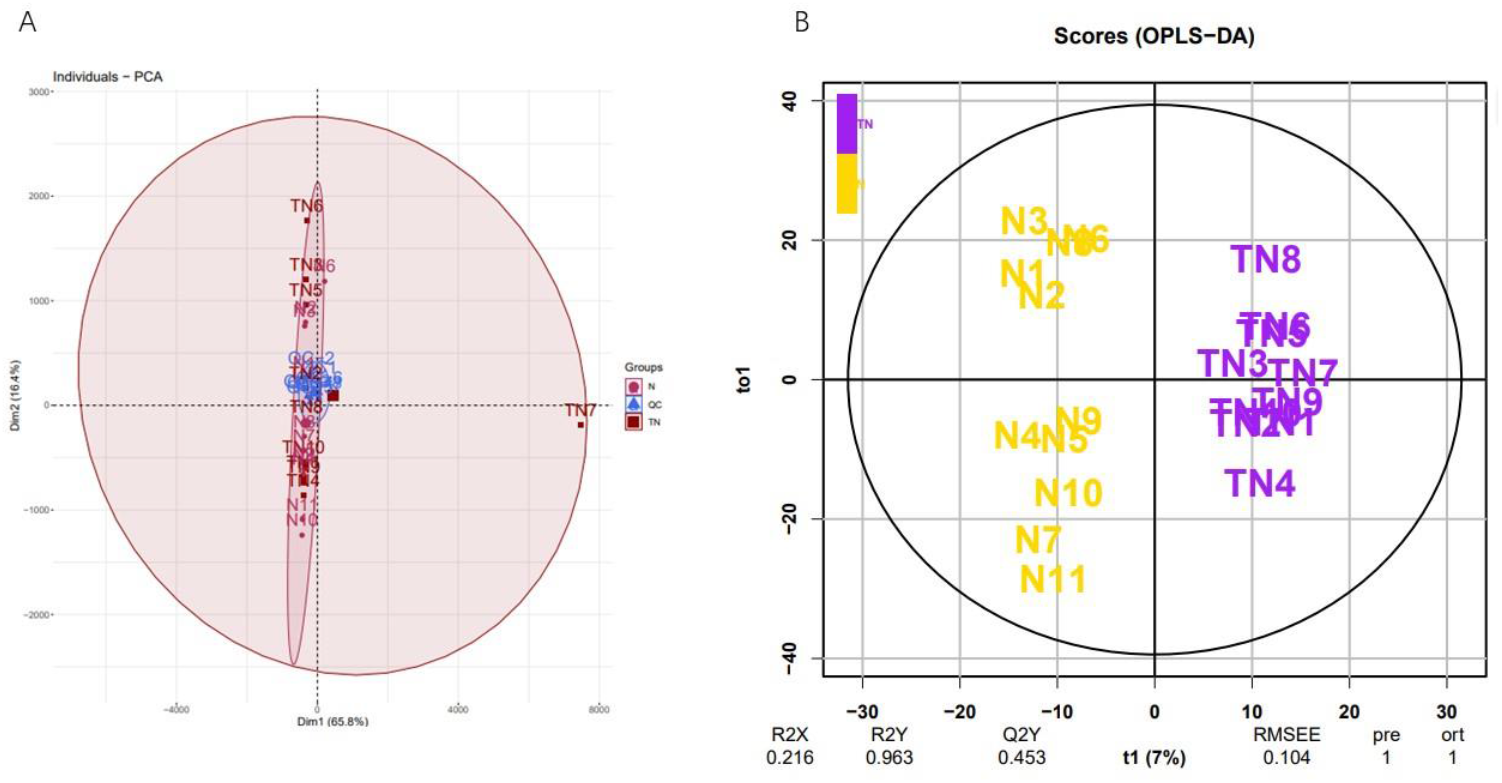
Visualized analysis of identified proteins in the three groups. A: principal component analysis (PCA); B: orthogonal partial least squares discrimination analysis (OPLS-DA) based on orthogonal signal correction.

### 3.2 Differential proteome analysis of whole urine proteome

#### 3.21 Quantitative analysis of differential proteins

The experimental samples of the two groups were compared between groups. Under loose screening condition, the screening criteria for differential proteins were: FC≥1.5 or≤0.67, two-tailed unpaired t-test P<0.05. Under strict conditions, the screening criteria were: FC≥2 or≤0.5, two-tailed unpaired t-test P<0.01. The number of differentially expressed proteins identified under different conditions is shown in Table 2. To further evaluate the reliability of differentially expressed proteins, the probability of random generation of differentially expressed proteins was calculated by random grouping. The total proteins identified by randomly selecting 11 healthy human samples and 10 TN patient samples were randomly grouped for verification (FC≥1.5 or≤0.67, P<0.05), and the average number of differentially expressed proteins generated was 64.2, indicating that at least 67.7% of the differentially expressed proteins were not generated randomly (Table 2); Random grouping verification was performed under more stringent conditions (FC≥2 or≤0.5, P<0.01), and the average number of differentially expressed proteins generated was 6.61, indicating that at least 87.1% of the differentially expressed proteins were not generated randomly (Table 2).

**Table 1.** shows the collected sample information and statistical results.

|  |  | TN<br>(n=10) | N<br>(n=11) |
| --- | --- | --- | --- |
| Gender | Male | 6 | 7 |
|  | Female | 4 | 4 |
| Age | Mean value | 53.9 | 55.8 |
|  | Standard deviation | 10.4 | 11.7 |

**Table 2.** Differential proteins of urine proteome.

| Differential protein screening conditions | number of differentially expressed proteins | Randomly generated number of different proteins | differential protein reliability |
| --- | --- | --- | --- |
| $FC \geq 1.5$ or $\leq 0.67$; $P < 0.05$ | 199 | 64.2 | 0.677 |
| $FC \geq 2$ or $\leq 0.5$; $P < 0.01$ | 51 | 6.6 | 0.871 |

Unsupervised clustering analysis of the whole protein of urine identification of TN and healthy controls, most of the urine proteome could separate patients with trigeminal neuralgia from healthy controls, there are still individual patients whose urine is close to that of healthy controls (Figure 3); It could be seen from the volcano map of urine total protein quantitative analysis that there are many proteins significant differences in the two groups.

**Figure 3.**
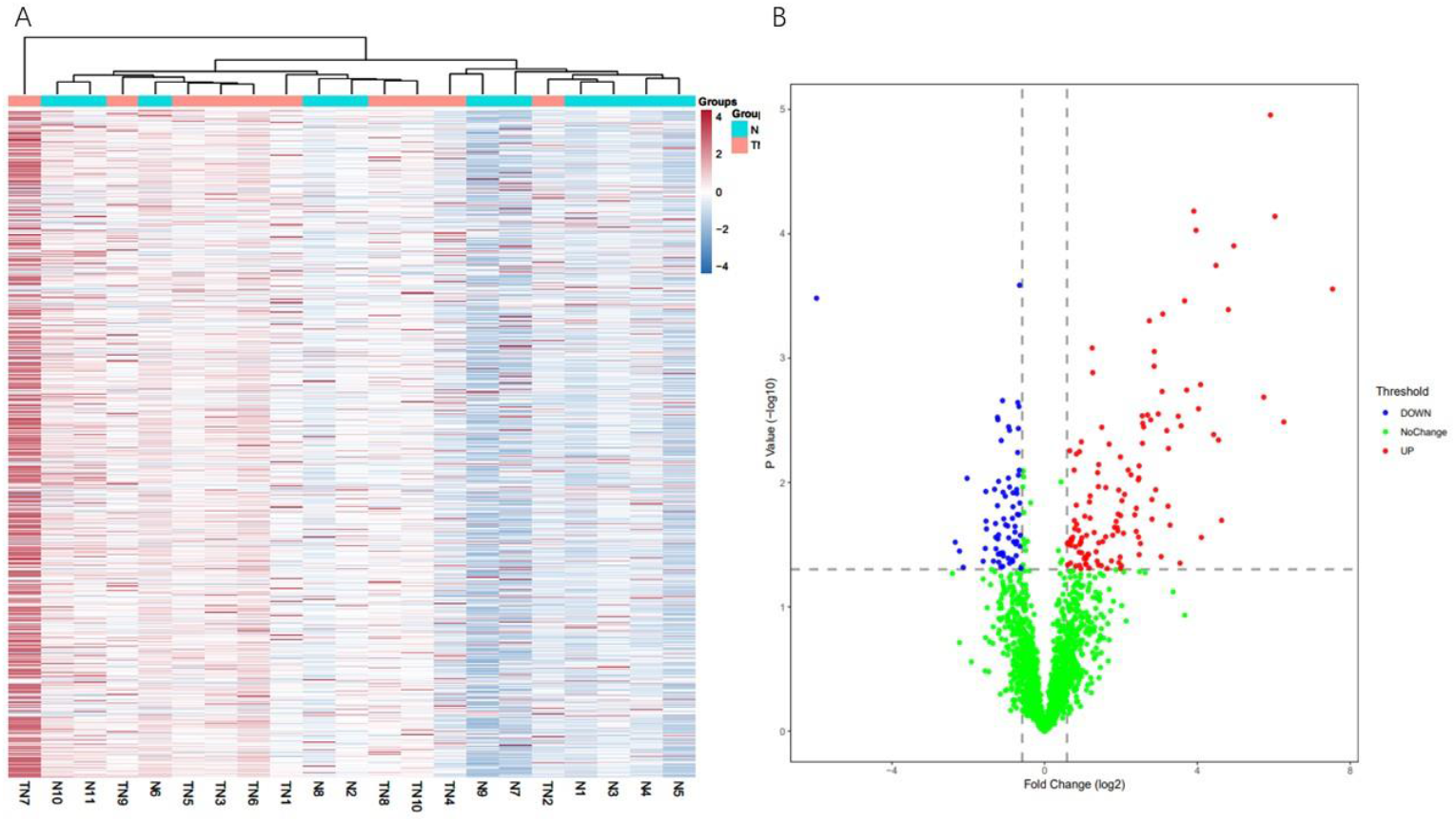
Unsupervised clustering analysis and volcano plot of urinary proteome in patients with TN and healthy controls

#### 3.22 Functional analysis of differentially expressed proteins

The reliability of differentially expressed proteins generated under strict screening conditions can reach more than 87% in randomized group experimental verification. In order to improve the reliability of the data, the functional analysis of differentially expressed proteins generated under strict screening conditions was selected. A total of 12 biological processes and 4 signal pathways were enriched (Table 3). Among the enriched biological processes, immune response regulatory signaling pathways, immune responses, and natural killer cell activation were significantly enriched. Among the enriched KEGG signaling pathways, antigen processing and presentation, natural killer cell-mediated cytotoxicity, and graft-versus-host disease were significantly enriched.

**Table 3.** GO functional enrichment analysis and KEGG pathway analysis of differentially expressed proteins.

| Category | Term | Count | % | P-Value |
| --- | --- | --- | --- | --- |
| GOTERM_BP_DIRECT | immune response-regulating signaling pathway | 11 | 16.9 | 2.30E-17 |
|  | immune response | 13 | 20 | 5.60E-08 |
|  | natural killer cell activation | 3 | 4.6 | 2.90E-03 |
|  | fatty acid transport | 3 | 4.6 | 4.10E-03 |
|  | natural killer cell inhibitory signaling pathway | 2 | 3.1 | 1.30E-02 |
|  | protein targeting to membrane | 3 | 4.6 | 1.30E-02 |
|  | positive regulation of gap junction assembly | 2 | 3.1 | 2.20E-02 |
|  | epidermis development | 3 | 4.6 | 2.80E-02 |
|  | positive regulation of CoA-transferase activity | 2 | 3.1 | 2.80E-02 |
|  | cytoplasmic translation | 3 | 4.6 | 3.80E-02 |
|  | epithelial cell differentiation | 3 | 4.6 | 3.90E-02 |
|  | monoatomic anion transport | 2 | 3.1 | 6.20E-02 |
| KEGG_PATHWAY | Antigen processing and presentation | 11 | 16.9 | 5.90E-13 |
|  | Natural killer cell mediated cytotoxicity | 11 | 16.9 | 8.30E-11 |
|  | Graft-versus-host disease | 6 | 9.2 | 9.40E-07 |
|  | Cholesterol metabolism | 3 | 4.6 | 1.90E-02 |

### 3.3 Individualized analysis of urine proteome

To understand the individual conditions of each patient with TN for individualized treatment, individual patient samples with 11 healthy group samples were compared one by one. The differentially expressed proteins identified under different conditions and their expression trends were shown in Table 4. There were a large number of significantly different proteins between the individual urine proteome of trigeminal neuralgia and healthy controls, which were significantly higher than the number of different proteins in the grouped protein analysis, further indicating that individual differences were widespread in the real world of medical care. Individualized analysis showed that there were significant differences between different patients and healthy controls. To discover as comprehensively as possible whether there were common differentially expressed proteins among patients with the disease, we performed a Venn diagram analysis of the differentially expressed proteins identified under relaxed conditions between trigeminal neuralgia and healthy controls. It was found that three differentially expressed proteins were identified in all 10 samples, nine proteins were identified in nine samples, and showed a relatively consistent expression change trend. Three proteins were identified simultaneously by nine proteins, but showed different change trends, as shown in Table 5.

**Table 4.** Number of identified proteins in individualized comparison of urine proteomics under different screening conditions

| Screening parameter | Trend of differential protein change | The number of significantly different proteins in individuals |  |  |  |  |  |  |  |  |  |
| --- | --- | --- | --- | --- | --- | --- | --- | --- | --- | --- | --- |
|  |  | 1 | 2 | 3 | 4 | 5 | 6 | 7 | 8 | 9 | 10 |
| P<0.05;<br>FC1.5 | total | 996 | 728 | 898 | 769 | 782 | 828 | 691 | 806 | 820 | 730 |
|  | ↑ | 604 | 319 | 362 | 429 | 355 | 365 | 392 | 463 | 506 | 442 |
|  | ↓ | 392 | 409 | 536 | 340 | 427 | 463 | 299 | 343 | 314 | 288 |
| P<0.01;<br>FC2 | total | 618 | 386 | 553 | 435 | 422 | 496 | 421 | 496 | 522 | 470 |
|  | ↑ | 396 | 164 | 221 | 278 | 202 | 209 | 263 | 326 | 366 | 295 |
|  | ↓ | 222 | 222 | 332 | 157 | 220 | 287 | 158 | 170 | 156 | 175 |

**Table 5.** List of proteins identified in urine by individualized analysis

| Protein ID | Protein Name | Variation trend | Proportion |
| --- | --- | --- | --- |
| Q9UJ70 | N-acetyl-D-glucosamine kinase (N-acetylglucosamine kinase) | ↑ | 9/10 |
| P00746 | Complement factor D | ↑ | 9/10 |
| P06727 | Apolipoprotein A-IV | ↑ | 9/10 |
| Q9HC38 | Glyoxalase domain-containing protein 4 | ↑ | 9/10 |
| P43490 | Nicotinamide phosphoribosyltransferase | ↑ | 9/10 |
| A0A075B6R9 | Probable non-functional immunoglobulin kappa variable | ↓ | 10/10 |
| Q86TX2 | Acyl-coenzyme A thioesterase 1 | ↓ | 10/10 |
| O60240 | Perilipin-1 | ↓ | 10/10 |
| Q8NE01 | Metal transporter CNNM3 | ↓ | 9/10 |
| Q9HAV5 | Tumor necrosis factor receptor superfamily member 27 | ↓ | 9/10 |
| Q9UHG0 | Doublecortin domain-containing protein 2 | ↓ | 9/10 |
| P01743 | Immunoglobulin heavy variable 1-46 | ↓ | 9/10 |
| P56385 | ATP synthase subunit e, mitochondrial | ↑8↓1 | 9/10 |
| P51159 | Ras-related protein Rab-27A | ↑7↓2 | 9/10 |
| Q96P63 | Serpin B12 | ↑5↓4 | 9/10 |

## 4 Experiment discussion

### 4.1 Urine proteome could well demonstrate the characteristics of TN patients

This study was the first to use urine proteome to study patients with TN. By visualizing the urine proteome of TN and healthy controls, it could be seen that the urine proteome can well distinguish different populations (Figure 2). Unsupervised clustering analysis of the urine whole proteome between the two groups showed that most of the samples in the same group were clustered together, and at the same time, it was seen that some individuals were similar to the samples between the other groups. This revealed that there are differences between individuals further. Due to the small number of specimens, no screening was performed on factors such as the severity of the disease, although this increased the complexity of the sample, it is more in line with the real situation of clinical diagnosis and treatment, and it just showed that the urine proteome has a unique advantage in the individualized analysis of patients.

### 4.2 Functional analysis of different proteins between groups

The screening of different proteins between groups was carried out under relaxed conditions and strict conditions respectively. In order to evaluate the accuracy of the screening results, the random grouping method was used in the study to calculate the possibility of random generation of differential proteins, and then to evaluate the possibility that the proteins were not randomly generated but were truly different from the samples. The data in table 2 showed that the overall reliability of the differential proteins was 87.1% under strict screening conditions. The random grouping method fully considered the differences between different sample individuals, and improved the reliability of the data.

In the enriched biological processes, immune response, natural killer cell activation, and natural killer cell inhibitory signaling pathways were all significantly enriched in comparison between trigeminal neuralgia patients and healthy controls. In KEGG analysis, the TN group and healthy control group were simultaneously enriched in signaling pathways such as antigen processing and presentation, natural killer cell-mediated cytotoxicity, and graft-versus-host disease, which were consistent with the enriched biological processes, indicating that the immune-related responses presented in the urine of patients were closely associated with trigeminal neuralgia. The infiltration of inflammatory cells and the release of cytokines such as IL-6 and TNF-α were considered key factors in initiating and maintaining pain ^8,9^. In EAE-induced female mice, adoptive transfer of Treg cells and spinal cord delivery of Treg cytokine interleukin-35 significantly reduced facial stimulation-induced pain and spontaneous pain ^10^. In this study, more than ten immune-related proteins were found, which contained 13 kinds of killer cell immunoglobulin-like receptors. Therefore, we speculated that killer cell-related immune factors might play an important role in the occurrence and development of trigeminal neuralgia. According to this mechanism, immune-related treatment of killer cells for patients with trigeminal neuralgia might be a worthy and potential treatment. The pathways of the immune system, axon guidance, and cellular stress response were related to the therapeutic effect of microvascular decompression ^11^, these response pathways were consistent with the urinary proteome, suggesting that the urinary proteome might also be an effective and non-invasive monitoring method for other treatments of trigeminal neuralgia.

### 4.3 Highly credible differential proteins in individual samples

In the experiment, one third or more of the total number of quantified proteins were identified as differentially expressed proteins under relaxed screening conditions (FC≥1.5 or≤0.67, two-tailed non-paired t-test P<0.05) through the analysis of individual samples and healthy control group, which was nearly twice as many as the differentially expressed proteins in group analysis, indicating that individual differences are common and have a great influence in the experiment. Therefore, it was more credible to find protein molecules with common characteristics through individualized analysis. A total of 15 highly credible protein molecules were identified in this experiment (up to 1 case was not identified in the sample of the group).

In the TN group, N-acetyl-D-glucosamine kinase,Complement factor D,Apolipoprotein A-IV, Glyoxalase domain-containing protein 4, Nicotinamide phosphoribosyltransferase significantly elevated. N-acetyl-D-glucosamine kinase participated in amino sugar metabolism, N-acetylneuraminic acid degradation and other pathways, N-acetylglucosamine kinase was highly expressed and plays a key role in the development of brain neuron dendrites ^12,13^, Urinary expression of complement factor D was increased, which was speculated to be related to nerve injury repair. Complement factor D was a serine protease of the alternative complement pathway and is essential for the formation of C3 convertase. It was a rate-limiting enzyme and might serve as a strategic target for advancing the therapeutic control of pathological complement activation ^14^, in this experiment, Complement factor D was found to be closely related to trigeminal neuralgia for the first time, and it may be involved in the occurrence of the disease, providing a new idea for disease-related immune treatment. Apolipoprotein A-IV was involved in numerous physiological processes, such as lipid absorption and metabolism, anti-atherosclerosis, platelet aggregation and thrombosis, glucose homeostasis and food intake ^15^, Apolipoprotein A-IV had been documented to significantly increase in perineural concentration in different animal models of sciatic nerve injury repair, and it was pointed out that it enters the nerve from the blood, and the article speculates that it may play a role in lipid transport in nerve tissue ^16^, our study found that it was significantly increased in TN, and it was speculated that there was the same biological process in trigeminal neuralgia. Glyoxalase domain-containing protein 4, whose gene family plays a wide role in metabolism, however, the role of this protein was little known ^17^, This study found that it was significantly elevated in TN, providing a direction for further study of the function of Glyoxalase domain-containing protein 4. Extracellular Nicotinamide phosphoribosyltransferas (NAMPT), also known as visfatin or pre-B cell colony-enhancing factor, had been shown to have adipocytokine, pro-inflammatory and pro-angiogenic activities, had the potential as a disease marker, and may be used for emerging extracellular NAMPT inhibitory therapies ^18^, at the same time, it was involved in promoting regenerative neurogenesis after ischemic stroke ^19^. In this experiment, 9/10 significant increases occurred in individualized analysis, and NAMPT was very likely to be involved in the disease process, providing a potential treatment direction for the disease.

Probable non-functional immunoglobulin kappa variable’ Acyl-coenzyme A thioesterase 1’Perilipin-1’Doublecortin domain-containing protein 2 ’Immunoglobulin heavy variable 1-46 were significantly decreased in TN patients. Doublecortin domain-containing protein 2 was widely distributed and highly expressed in the brain, and inhibited the classical Wnt signaling pathway ^20^, and it was also related to reading ability and the development of regulatory nerves ^21^, in our experiment, 9/10 showed a reduction, which was very likely to be related to nerve damage. Immunoglobulin heavy variable 1-46, participated in antigen recognition in immune response ^22^, 9/10 showed a reduction; Perilipin-1, Probable non-functional immunoglobulin kappa variable and Acyl-coenzyme A thioesterase 1 showed significant reductions; although there have been no reports of association with neuropathy, we speculated that it may be associated with the occurrence of neuropathy, this study provided clues for further study of neuropathy. Metal transporter CNNM3 Participate in metal ion transport, reduced in TN patients, and there was no relevant report on its related mechanisms. Tumor necrosis factor receptor superfamily member 27 Tumor necrosis factor, as a pro-inflammatory cytokine, can enhance the development of pain and hyperalgesia by mediating the activation of NF-kappa-B and JNK pathways^23^, our study found that its receptor was reduced, and whether it is a protective mechanism of the body needs further verification.

Meanwhile, we also focused on some protein molecules that significantly changed in the grouped analysis. Discoidin, CUB and LCCL domain-containing 2 (DCBLD2) is a neurofibrin-like transmembrane scaffold receptor with known and expected roles in vascular remodeling and neuronal positioning, and was also upregulated in tumors ^24^, this high-confidence molecules were most likely to become disease-related markers. In addition, some of the co-identified proteins in the individualized analysis showed different or even opposite changes in different cases (Table 5), such as ATP synthase subunit e, mitochondrial, Ras-related protein Rab-27A and Serpin B12. ATP synthase subunit e, mitochondrial was a multifunctional enzyme complex involved in ATP generation, which was significantly different in different TN patients. It has been reported in the literature that adenosine triphosphate (ATP) is a key regulator in the nociceptive pathway. The release of ATP from damaged tissues or sympathetic efferent nerves sensitizes peripheral sensory neurons and enhances downstream central sensitization mechanisms, a situation considered to be the basis of many chronic pain conditions. Preclinical use of purinergic receptor P2×3 and P2×2/3 antagonists on sensory nerves responsible for ATP signaling can improve pain^25^. If so, urinary ATP synthase subunit e, mitochondrial may provide guidance for the use of purinergic receptor P2×3 and P2×2/3 antagonists in the treatment of pain. Ras-related protein Rab-27A is a small GTPase that cycles between the active GTP-bound state and the inactive GDP-bound state. In its active state, it binded to a variety of effector proteins to regulate the homeostasis of the late endocytic pathway, including endosomal localization, maturation, and secretion ^26^, Meanwhile, it also played a role in the secretion of cytotoxic granules in lymphocytes ^27^. Serpin B12 inhibits trypsin and plasmin, but does not inhibit thrombin, factor Xa, or urokinase-type plasminogen activator ^28^. The significant differences in these proteins between different patients merit further study.

### 4.4 Problems and prospects

This study was the first to use proteomic techniques to study the urine of patients with TN. Although the data were analyzed from different angles to ensure the reliability of the experimental results, the number of specimens studied was relatively small and it was not possible to exhaustively study the commonalities and differences between different cases. In the future, a multi-center large sample size study of TN will have a more profound impact. The study found that the immune response was strong in TN patients, especially in the identification of 13 killer cell immunoglobulin-like receptors, but there is clearly insufficient immune-related treatment for TN at present, which deserves the attention of doctors and extensive research.

## 5 Conclusions

This is the first study to comprehensively profile TN using urinary proteomics, deeply describing the common and differential characteristics of TN patients, and providing important clues for disease pathogenesis and treatment. At the same time, high-confidence proteins related to disease were identified in the experiment, which may become disease markers to provide help for disease diagnosis and treatment.

## Supporting information

Supplementary data for Clinical information and DIFFERENT PROTEIN

## Data Availability

All data produced in the present study are available upon reasonable request to the authors

## Authors’ contributions

Lilong Wei: project administration, writing – original draft preparation. Haitong Wang: investigation, formal analysis. Yun Zhou: specimen collection. Jianqiang Wu: mass spectrometry analysis. Yuliang Zhan: methodology. Yongtong Cao: methodology, funding acquisition. Youhe Gao: conceptualization, writing – review & editing.

Lilong Wei and Haitong Wang contributed equally to this study. Youhe Gao, Yuliang Zhan and Yongtong Cao are corresponding authors. All authors have checked and approved the final version of the manuscript.

## Data availability statement

The data that supports the findings of this study are available in the supplementary material of this article.

## Ethics approval statement

The study was approved by the Ethics Committee of China-Japan Friendship Hospital (approval number: 2023-KY-126) and implemented in strict accordance with the relevant ethics standards and Declaration of Helsinki.

## Consent statement

The study collected the remaining specimens after clinical testing and conducted a retrospective study, excluding personal privacy. The ethics committee has approved the application for exemption from informed consent. The manuscript does not contain data from any individual person; therefore, participant consent for publication is not applicable.

## References

1 Carnevale, J. A. & Knopman, J. Atrophy and Severe Kinking of Trigeminal Nerve Root by Duplicate Trunks of Superior Cerebellar Artery in an Elderly Patient with Trigeminal Neuralgia. World Neurosurg 179, 100–101 (2023).

2 Headache Classification Committee of the International Headache Society (IHS) The International Classification of Headache Disorders, 3rd edition. Cephalalgia 38, 1–211 (2018).

3 Finnerup, N. B., Kuner, R. & Jensen, T. S. Neuropathic Pain: From Mechanisms to Treatment. Physiol Rev 101, 259–301 (2021).

4 Gallien, S. & Domon, B. Advances in high-resolution quantitative proteomics: implications for clinical applications. Expert Rev Proteomics 12, 489–498 (2015).

5 Dayon, L., Cominetti, O. & Affolter, M. Proteomics of human biological fluids for biomarker discoveries: technical advances and recent applications. Expert Rev Proteomics 19, 131–151 (2022).

6 Shao, C., Wang, Y. & Gao, Y. Applications of urinary proteomics in biomarker discovery. Sci China Life Sci 54, 409–417 (2011).

7 高友鹤. 尿液有可能成为生物标志物的金矿吗? 中国科学 生命科学 (2013).

8 Balkowiec-Iskra, E. [The role of immune system in inflammatory pain pathophysiology]. Pol Merkur Lekarski 29, 395–399 (2010).

9 Iuodzhbalis, G. & Sabalis, G. I. [The immune status of patients with trigeminal neuralgia and its correction]. Stomatologiia (Mosk) 68, 44–46 (1989).

10 Duffy, S. S. et al. Regulatory T Cells and Their Derived Cytokine, Interleukin-35, Reduce Pain in Experimental Autoimmune Encephalomyelitis. J Neurosci 39, 2326–2346 (2019).

11 Doshi, T. L., Dorsey, S. G., Huang, W., Kane, M. A. & Lim, M. Proteomic Analysis to Identify Prospective Biomarkers of Treatment Outcome After Microvascular Decompression for Trigeminal Neuralgia: A Preliminary Study. J Pain 25, 781–790 (2024).

12 Lee, H., Dutta, S. & Moon, I. S. Upregulation of dendritic arborization by N-acetyl-D-glucosamine kinase is not dependent on its kinase activity. Mol Cells 37, 322–329 (2014).

13 Islam, M. A., Sharif, S. R., Lee, H. & Moon, I. S. N-Acetyl-D-Glucosamine Kinase Promotes the Axonal Growth of Developing Neurons. Mol Cells 38, 876–885 (2015).

14 Barratt, J. & Weitz, I. Complement Factor D as a Strategic Target for Regulating the Alternative Complement Pathway. Front Immunol 12, 712572 (2021).

15 Qu, J., Ko, C. W., Tso, P. & Bhargava, A. Apolipoprotein A-IV: A Multifunctional Protein Involved in Protection against Atherosclerosis and Diabetes. Cells 8 (2019).

16 Boyles, J. K., Notterpek, L. M. & Anderson, L. J. Accumulation of apolipoproteins in the regenerating and remyelinating mammalian peripheral nerve. Identification of apolipoprotein D, apolipoprotein A-IV, apolipoprotein E, and apolipoprotein A-I. J Biol Chem 265, 17805–17815 (1990).

17 Farrera, D. O. & Galligan, J. J. The Human Glyoxalase Gene Family in Health and Disease. Chem Res Toxicol 35, 1766–1776 (2022).

18 Semerena, E., Nencioni, A. & Masternak, K. Extracellular nicotinamide phosphoribosyltransferase: role in disease pathophysiology and as a biomarker. Front Immunol 14, 1268756 (2023).

19 Zhao, Y. et al. Regenerative Neurogenesis After Ischemic Stroke Promoted by Nicotinamide Phosphoribosyltransferase-Nicotinamide Adenine Dinucleotide Cascade. Stroke 46, 1966–1974 (2015).

20 Schueler, M. et al. DCDC2 mutations cause a renal-hepatic ciliopathy by disrupting Wnt signaling. Am J Hum Genet 96, 81–92 (2015).

21 Meng, H. et al. DCDC2 is associated with reading disability and modulates neuronal development in the brain. Proc Natl Acad Sci U S A 102, 17053–17058 (2005).

22 Lefranc, M. P. Immunoglobulin and T Cell Receptor Genes: IMGT((R)) and the Birth and Rise of Immunoinformatics. Front Immunol 5, 22 (2014).

23 Khan, A. A. et al. Tumor necrosis factor alpha enhances the sensitivity of rat trigeminal neurons to capsaicin. Neuroscience 155, 503–509 (2008).

24 Schmoker, A. M. et al. Dynamic multi-site phosphorylation by Fyn and Abl drives the interaction between CRKL and the novel scaffolding receptors DCBLD1 and DCBLD2. Biochem J 474, 3963–3984 (2017).

25 Krajewski, J. L. P2×3-Containing Receptors as Targets for the Treatment of Chronic Pain. Neurotherapeutics 17, 826–838 (2020).

26 Zhang, J. et al. DENN domain-containing protein FAM45A regulates the homeostasis of late/multivesicular endosomes. Biochim Biophys Acta Mol Cell Res 1866, 916–929 (2019).

27 Menasche, G. et al. A newly identified isoform of Slp2a associates with Rab27a in cytotoxic T cells and participates to cytotoxic granule secretion. Blood 112, 5052–5062 (2008).

28 Askew, Y. S. et al. SERPINB12 is a novel member of the human ov-serpin family that is widely expressed and inhibits trypsin-like serine proteinases. J Biol Chem 276, 49320–49330 (2001).

